# Association of leucocyte telomere length with frailty: a large–scale cross–sectional analysis in UK Biobank

**DOI:** 10.1101/2021.06.11.21258736

**Authors:** Vasiliki Bountziouka, Christopher P Nelson, Veryan Codd, Qingning Wang, Crispin Musicha, Elias Allara, Stephen Kaptoge, Emanuele Di Angelantonio, Adam S Butterworth, John R Thompson, Elizabeth M Curtis, Angela M Wood, John N Danesh, Nicholas C Harvey, Cyrus Cooper, Nilesh J Samani

## Abstract

**Background:** Leucocyte telomere length (LTL), a potential marker of biological age, has been associated with risk of many diseases. We investigated whether LTL is associated with risk of frailty, a multidimensional syndrome of decline that affects multiple systems and predisposes to adverse health outcomes.

**Methods:** In a cross–sectional analysis, we studied 441,781 UK Biobank participants (aged 40–70 years), with complete data on LTL and frailty indicators. We defined frailty as the presence of at least three of five indicators: weaker grip strength, slower walking pace, weight loss in the past year, lower physical activity, and exhaustion in the past two weeks. We evaluated association of LTL with frailty using adjusted (chronological age, sex, deprivation, smoking, alcohol intake, body mass index, multimorbidity) multinomial and ordinal regression models. We used Mendelian randomisation (MR), using 131 genetic variants associated with LTL, to assess if the association of LTL with frailty was causal.

**Findings:** Frail participants (4·6%) were older (median age difference (95% CI): 3 (2·5; 3·5) years), more likely to be female (61%), and had shorter LTL (−0·13SD vs 0·03SD) than non–frail. In adjusted analyses, both age and LTL were associated with frailty (OR=1·03 (95%CI: 1·02–1·04) per year of older chronological age; 1·10 (1·08; 1·11) per SD shorter LTL). Within each age group (40–49, 50–59, 60–69 years) the prevalence of frailty was about 33% higher in participants with shorter (−2SD) versus longer telomeres (+2SD). MR analysis showed an association of LTL with frailty that was directionally consistent with the observational association, but not statistically significant.

**Interpretation:** Inter–individual variation in LTL is associated with the risk of frailty independently of chronological age and other risk factors. Our findings provide evidence for an additional biological determinant of frailty.

**Funding:** The UK Medical Research Council, the Biotechnology and Biological Sciences Research Council and the British Heart Foundation (MRC grant: MR/M012816/1) funded our measurements of LTL in UK Biobank. The funders had no role in study design; the collection, analysis, and interpretation of data; the writing of the report; and the decision to submit the paper for publication.

**Research in context:** *Evidence before this study:* Telomere length has been proposed as a biomarker of biological age. Shorter leucocyte telomere length (LTL) is associated with higher incidence of several age–associated diseases, spanning multiple body systems, and with lower life expectancy. However, the association of shorter LTL with frailty, a multidimensional syndrome of decline across multiple systems, is inconclusive. We searched PubMed using the terms “telomere length AND frailty” in the title, abstract, or text with no language restrictions. This search identified 50 papers published before 30 November, 2020. Of these, three systematic reviews and/ or meta–analyses of modest size (five to nine studies, with total n<10,000) and 11 original research articles reported on associations between telomere length and frailty. These small–scale studies, primarily in older individuals, do not support an independent association between shorter LTL and higher risk of frailty.

*Added value of this study:* Our large–scale population–based study, involving over 441,000 participants, provides strong evidence that shorter LTL is associated with higher risk of both pre–frailty and frailty even after adjustment for chronological age and other established determinants of frailty, namely, age, sex, alcohol intake, smoking, obesity, deprivation and multimorbidity. Within each age group (40–49, 50–59, 60–69 years) the prevalence of frailty was about 33% higher in participants with shorter (−2SD) versus longer telomeres (+2SD). Part of the association of shorter LTL with higher risk of frailty may be due to the association in aggregate of LTL with diseases. Findings using genetic variants associated with LTL as instruments were directionally consistent with the observational association, but not statistically significant.

*Implications of all the available evidence:* Shorter LTL is associated with frailty independent of chronological age and several other risk factors. The observation that risk of frailty associated with shorter LTL is proportionately similar across the age range of 40–70 years suggests that shorter LTL acts through a mechanism beyond just accelerating the impact of chronological ageing on risk of frailty.

## Introduction

Frailty is a multidimensional syndrome of decline that affects multiple systems and predisposes to adverse health outcomes.^1^ It is associated with greater vulnerability to stressors and increased risk of adverse health outcomes, including falls, fractures, hospitalisation and death.^2^ Frailty is inter–related, but not synonymous, with comorbidity and disability. Approaches to its operational definition include an accumulation of deficits as proposed by Rockwood et al.,^1^ or a specific biological syndrome, characterised by weight loss, fatigue, reduced muscle strength, reduced walking speed and low physical activity, as proposed by Fried.^3^ Both definitions have strengths and weaknesses: the Fried physical frailty phenotype includes two components of sarcopenia and may therefore overlap considerably with muscle function; the Rockwood approach essentially assesses the number of comorbidities, with its attendant circularity of cause and effect. Attempts have been made to achieve consensus on the definition of frailty, recognising that it is characterized by a plethora of physical, psychological, physiological, and social life aspects that co–exist in complex combinations.^4–7^

Although frailty is more prevalent in older people, it does not occur exclusively above any specific chronological age threshold.^8^ Hence, there is a need to identify other biological factors that may predispose to frailty. There is a particular interest in whether biological age, as distinct from chronological age, is associated with risk of frailty.^9^ Telomere length has emerged as a potential biomarker of biological age, with shorter telomeres indicating more advanced biological age.^10^ Shorter mean leucocyte telomere length (LTL) has been associated with risk of several age–associated diseases with causal inference analyses suggesting that some of the associations are primary.^11,12^ However, it should be noted that the relationship between LTL and disease is complex and longer LTL can also be associated with disease risk, most notably for several cancers.^11–13^ Current evidence on whether inter–individual variation in LTL is associated with higher risk of frailty is inconclusive.^14–16^

We have recently generated cohort–wide LTL measurements in UK Biobank (UKB).^17^ Using this large–scale resource we investigated whether LTL is associated with frailty independently of chronological age and other established risk factors.

## Methods

### Participants and Data Collection

As previously described,^18^ UKB recruited 502,478 participants aged 40–69 years during the years 2006–2010. Participants have been characterised in detail using questionnaires, physical measurements, biological assays and longitudinal linkage with multiple health record systems. Detailed information regarding the physical assessments undertaken is available at https://www.ukbiobank.ac.uk/. UKB received approval from the North West Centre for Research Ethics Committee (11/NW/0382). The use of data presented in this paper was approved by the Access Committee of UKB under application number 6077.

### Frailty phenotype

Based on the concept of frailty as a biological syndrome, we implemented the ‘*phenotype*’ model of frailty, developed by Fried et al.,^3^ which has been previously utilised in the UKB.^19,20^ Under this model, five indicators, assessed at baseline examination, were used to define frailty: weakness, slowness, weight loss, low physical activity and exhaustion. For each indicator we employed the following scoring system: (i) weakness, measured using the maximum hand grip strength from both arms (field codes: “46” and “47”): participants in the lowest 20% of the cohort (sex and body mass index (BMI) adjusted) were considered to meet the frailty criteria and thus given a score of “1” or “0” otherwise; (ii) slowness, measured using the self–reported walking pace (field code: “924”): “1” for slow pace, “0” otherwise; (iii) weight loss, measured through the self–reported weight change (field code: “2306”) compared with one year ago: “1” for yes–weight loss, “0” otherwise; (iv) low physical activity, measured through self–reported types of physical activity (field code: “6164”) in the last 4 weeks: “1” for non or light activity (e.g. pruning, watering the lawn), “0” otherwise (e.g. weeding, lawn mowing, carpentry and digging, walking for pleasure, swimming, cycling, or other strenuous sports), and v) exhaustion, measured through self–reported tiredness/ lethargy in last 2 weeks (field code: “2080”): “1” for more than half the days or nearly every day, “0” otherwise.

Participants who responded “Don’t know” or “Prefer not to answer” to any of the five frailty indicators, or with missing values, were excluded from the analysis. People with one or two indicators in aggregate were classified as pre–frail, whilst frailty was defined as the presence of three or more of the five indicators.

### Leucocyte telomere length (LTL) measurement

LTL measurements were undertaken on DNA collected at baseline and quality controlled as described elsewhere.^17^ Briefly, LTL was measured using a validated qPCR method and reported as a ratio of the telomere repeat number (T) to a single–copy gene (S) (T/S ratio). The measurements were loge–transformed to approximate the normal distribution. We utilised z–standardised values of LTL (field code: “22192”) to facilitate comparison with other datasets.^17^

### Other phenotypes

To adjust for other known or potential determinants of frailty,^3^ we extracted information on the following phenotypes also collected at baseline: Social deprivation score (based on quintiles of Townsend Index deprivation score at the time of recruitment (field code: “189”), derived from the 2011 Census UK data,^21^ with 1st quintile being most affluent), smoking (self–reported field code “20116” and classified as non–smoker; ex–smoker; current smoker), alcohol intake (self–reported frequency of alcohol intake (field code “1558”) and classified as never/ special occasions only; 1–3 times per month; 1–4 times per week; daily/ almost daily), body mass index (field code “21001” categorised as underweight (<18·5 kg/m^2^); normal weight (18·5–24·9 kg/m^2^); overweight (25–29·9 kg/m^2^); obese (≥30 kg/m^2^)) and multimorbidity,^22^ measured as the total number of additional long–term medical conditions (LTC; pooled out from the self–reported non–cancer illness code (field code “20002”) and the cancer diagnosed by doctor code (field code “2453”)) and classified as none; one LTC; two LTC; three LTC; four or more LTC.

### Statistical analysis

Descriptive statistics are shown as mean (SD), median (1st quartile, 3rd quartile) or frequencies (%). For primary analysis we used multinomial logistic regression models to investigate the association of chronological age and LTL with frailty status, defined as non–frail, pre–frail and frail. Interaction and quadratic terms for age, LTL and sex were tested and the model with the lowest Bayesian information criterion (BIC) was selected. The best model was then adjusted for other potential determinants of frailty. Results are shown as relative risk ratios (RRR) along with their corresponding 95% confidence intervals (95% CI). The average adjusted prediction for the frequency of frailty was plotted against chronological age and LTL. We also report observed associations of the average (“usual”) LTL values, adjusted for the regression dilution ratio (RDR) of 0·68 (0·64–0·72) for loge–LTL that was derived using 1351 serial measurements of LTL taken at mean interval of 5.5 years (range: 2–10 years).^17^ Secondary analysis involved multinomial regression and binary logistic regression to assess associations (RRR and odds ratios (OR)) with the number of frailty indicators and the individual frailty indicators respectively.

We have previously characterised the association of LTL with 123 diseases, identified using hospital admissions, operations, death registry and self–report data as described elsewhere.^12^ To assess the extent to which any association of LTL with frailty is independent of any association of LTL with these diseases at baseline, we conducted regression modelling using standardised residuals after regressing LTL on indicators of history of the 123 diseases.

To investigate whether any relationship between LTL and frailty is causal, we conducted Mendelian randomisation (MR) analyses,^23^ using 131 independent and uncorrelated genetic variants associated with LTL at genome–wide significance^12^ as instrumental variables. Further details for the statistical analysis, including the MR analysis, are provided in the “*Methods section*” of the Supplementary material.

## Results

Of 472,174 participants in UK Biobank with a valid LTL measurement, we excluded 30,393 (6·4%) from the current analysis because they lacked information on frailty indicators, relevant covariates, or both (**Supplementary Figure 1**). There was no difference in the distribution of sex and age between participants who were included or excluded from the analysis (females 54·2% vs 54·6%; mean age 56·5 vs 57, respectively). However, those who were excluded had on average shorter telomeres compared to the complete cases for the analysis (−0·019SD vs 0·001SD).

Of the 441,781 participants included in the analysis, 223,648 (51%) had no frailty indicators, 147,789 (33%) had one indicator, 49,826 (11%) had two indicators, 15,387 (3·5%) had three indicators, 4,473 (1%) had four indicators, and 658 (0·15%) had all five indicators. Hence, 20,518 (4·6%) participants met the criteria for frailty, and 197,615 (44·7%) for pre–frailty (**Table 1**). Compared to non–frail participants, frail participants were older and more likely to be female, socioeconomically deprived, current smokers, obese, alcohol drinkers and report multiple LTCs (**Table 1**). Compared with non–frail participants mean LTL was shorter in both frail and in pre–frail participants, with a greater magnitude of difference for frail participants (**Table 1**).

**Table 1:** Distribution of demographic and clinical characteristics and leucocyte telomere length (LTL), overall and across participants’ frailty status.

|  | Frailty status |  |  |
| --- | --- | --- | --- |
|  | Non-frail | Pre-frail | Frail |
|  |  | (1–2 indicators) | (3–5 indicators) |
| n (%) | 223,648 (51) | 197,615 (45) | 20,518 (4.6) |
| Age, years | 57 (49; 62) | 59 (51; 64) | 60 (53; 64) |
| Females, n (%) | 115,835 (52) | 111,119 (56) | 12,436 (61) |
| LTL, SD | 0.03 (0.99) | -0.02 (1.00) | -0.13 (1.00) |
| Fifths of deprivation, n (%) |  |  |  |
| 1st (least deprived) | 87,138 (39) | 63,773 (32) | 4,084 (20) |
| 2nd | 50,480 (23) | 41,727 (21) | 3,383 (16) |
| 3rd | 36,698 (16) | 33,617 (17) | 3,327 (16) |
| 4th | 30,047 (13) | 31,990 (16) | 4,389 (21) |
| 5th (most deprived) | 19,285 (8.6) | 26,508 (13) | 5,335 (26) |
| Smoking status, n (%) |  |  |  |
| Never | 126,833 (57) | 105,566 (53) | 9,264 (45) |
| Previous | 77,617 (35) | 69,432 (35) | 7,172 (35) |
| Current | 19,198 (8.6) | 22,617 (11) | 4,082 (20) |
| Frequency of alcohol intake, n (%) |  |  |  |
| Daily | 51,759 (23) | 37,374 (19) | 2,484 (12) |
| 1–4 times/week | 118,555 (53) | 92,804 (47) | 6,745 (33) |
| 1–3 times/month | 23,015 (10) | 23,627 (12) | 2,461 (12) |
| Occasionally/ never | 30,319 (14) | 43,810 (22) | 8,828 (43) |
| Categories of body mass index, n (%) |  |  |  |
| <18 kg/m<sup>2</sup> | 1,071 (0.48) | 992 (0.50) | 160 (0.78) |
| 18–25 kg/m<sup>2</sup> | 81,510 (36) | 58,962 (30) | 4,138 (20) |
| 25–30 kg/m<sup>2</sup> | 98,727 (44) | 82,959 (42) | 6,707 (33) |
| ≥30 kg/m<sup>2</sup> | 42,340 (19) | 54,702 (28) | 9,513 (46) |
| Number of comorbidities, n (%) |  |  |  |
| None | 66,333 (30) | 39,732 (20) | 1,307 (6.4) |
| One long term condition | 65,376 (29) | 50,320 (25) | 2,690 (13) |
| Two long term conditions | 44,089 (20) | 41,618 (21) | 3,483 (17) |
| Three long term conditions | 24,672 (11) | 28,595 (14) | 3,563 (17) |
| Four or more long term conditions | 23,178 (10) | 37,350 (19) | 9,475 (46) |

Results shown as mean (SD) or median (1<sup>st</sup> quartile; 3<sup>rd</sup> quartile), unless otherwise indicated. Leucocyte telomere length (LTL) measurements are z-standardised.

Shorter LTL was associated with higher odds of having each of the individual frailty indicators (**Table 2)**. Similarly, when participants were dichotomised by the number of frailty indicators, we observed that shorter LTL was associated with higher relative risk of having greater number of frailty indicators (**Supplementary Table 1**). In analyses subdivided by age and sex (**Table 3**), we found higher prevalence of pre–frailty and frailty at older ages for both men and women. Mean LTL was higher in women than men, lower at older ages for both sexes, and declined with frailty across all age groups (**Table 3**).

**Table 2:** Adjusted odds ratios (OR) and 95% confidence intervals (95% CI) from a binary logistic regression model^∫^ of leucocyte telomere length on frailty indicators.

|  | n (%) | OR | (95% CI) | P |
| --- | --- | --- | --- | --- |
| *Weakness (hand grip strength)* | 101,179 (23) |  |  |  |
| Telomere length, per SD shorter |  | 1.02 | (1.01; 1.03) | <0.0001 |
| *Slowness (walking pace)* | 34,421 (7.8) |  |  |  |
| Telomere length, per SD shorter |  | 1.06 | (1.05; 1.07) | <0.0001 |
| *Weight loss* | 67,684 (15) |  |  |  |
| Telomere length, per SD shorter |  | 1.02 | (1.01; 1.03) | <0.0001 |
| *Low physical activity* | 56,805 (13) |  |  |  |
| Telomere length, per SD shorter |  | 1.04 | (1.03; 1.05) | <0.0001 |
| *Exhaustion (tiredness/ lethargy)* | 54,695 (12) |  |  |  |
| Telomere length, per SD shorter |  | 1.03 | (1.02; 1.04) | <0.0001 |

<sup>†</sup>Models additionally adjusted for age, sex, quintiles of Townsend index of deprivation (2011), smoking, alcohol intake, body mass index and number of long-term medical conditions.

**Table 3:** Age and leucocyte telomere length (LTL) distribution between males and females across their frailty status.

|  | Frailty status |  |  |
| --- | --- | --- | --- |
|  | Non-frail | Pre-frail | Frail |
|  |  | (1–2 indicators) | (3–5 indicators) |
| *Females* |  |  |  |
| n (%) | 115,835 (48) | 111,119 (46) | 12,436 (5.2) |
| Age, years | 56 (49; 62) | 58 (51; 63) | 59 (52; 64) |
| 40 to 49, n (%) | 30,965 (54) | 23,783 (42) | 2,140 (3.8) |
| 50 to 59, n (%) | 41,018 (50) | 37,231 (45) | 4,427 (5.4) |
| 60 to 70, n (%) | 43,852 (44) | 50,105 (50) | 5,869 (5.9) |
| LTL, SD |  |  |  |
| Overall | 0.12 (0.98) | 0.06 (0.99) | -0.03 (0.99) |
| 40 to 49 | 0.32 (0.97) | 0.31 (0.99) | 0.26 (1.00) |
| 50 to 59 | 0.16 (0.97) | 0.13 (0.98) | 0.02 (0.95) |
| 60 to 70 | -0.06 (0.97) | -0.10 (0.97) | -0.19 (0.99) |
| *Males* |  |  |  |
| n (%) | 107,813 (53) | 86,496 (43) | 8,082 (4.0) |
| Age, years | 57 (49; 63) | 59 (51; 64) | 60 (54; 65) |
| 40 to 49, n (%) | 26,964 (58) | 18,571 (40) | 1,202 (2.6) |
| 50 to 59, n (%) | 35,797 (55) | 26,488 (41) | 2,462 (3.8) |
| 60 to 70, n (%) | 45,052 (50) | 41,437 (46) | 4,418 (4.9) |
| LTL, SD |  |  |  |
| Overall | -0.07 (0.98) | -0.12 (0.99) | -0.28 (1.01) |
| 40 to 49 | 0.22 (0.96) | 0.21 (0.97) | 0.11 (0.96) |
| 50 to 59 | -0.03 (0.96) | -0.05 (0.97) | -0.19 (0.97) |
| 60 to 70 | -0.27 (0.96) | -0.32 (0.97) | -0.43 (1.01) |

Results shown as mean (SD) or median (1<sup>st</sup> quartile; 3<sup>rd</sup> quartile), unless otherwise indicated. Leucocyte telomere length (LTL) measurements are z-standardised.

To assess the relationships between age, LTL and frailty, a model with linear and quadratic terms for age and LTL was found to be the best model minimizing BIC (model M5, **Supplementary Table 2**). In the presence of the quadratic terms, the age*LTL interaction was non–significant (**Supplementary Table 2**). Furthermore, the fitted values for the prevalence of non–frailty, pre–frailty and frailty obtained from M5, for the average LTL (i.e. z–LTL=0SD), were similar to the observed frequencies with the small caveat that a crossover between the distributions of non–frailty and pre–frailty in the model occurs a year earlier (**Supplementary Figure 2**).

Age was positively associated with being frail compared to being non–frail with an average 3·4% higher risk per year of chronological age (M5; **Supplementary Table 2**). Similarly, shorter LTL was positively associated with higher risk of frailty (11.5% higher risk for one SD shorter LTL), with the rate of change in the association of LTL with frailty dependent on the length of the telomere and modestly greater for shorter telomeres (p for quadratic term=0·001) (**Supplementary Table 2**). There were analogous associations of age and LTL with pre–frailty, but weaker than with frailty (**Supplementary Table 2**). The coefficients for age and LTL from M5 were also similar to the ones derived from a generalised ordinal model (**Supplementary Table 3**).

We next evaluated whether sex or the presence of other factors associated with frailty shown in **Table 1**, impacted on the effects of age and LTL on risk of frailty. Adjustment for these factors did not substantially alter the association of age with frailty (3·2% higher risk per year of chronological age, **Table 4**). Similarly, the association of shorter LTL with pre–frailty (2·1% (1·4%; 2·7%) higher relative risk per one SD shorter LTL) and frailty (9·6% (7·9%; 11%) higher relative risk per one SD shorter LTL) remained highly significant after adjustment for these factors, with the rate of change of the association again modestly dependent on the length of the telomere (p for LTL quadratic term: 0·003 for pre–frailty and 0·03 for frailty) (**Table 4**). The relative risk of frailty with different LTLs in men and women at age 40, 55 and 70, standardised to a 40 year old male, is shown in **Figure 1**. Across the spectrum of possible situations there was a greater than three–fold difference in the relative risk of frailty associated with variation in LTL.

**Figure 1:**
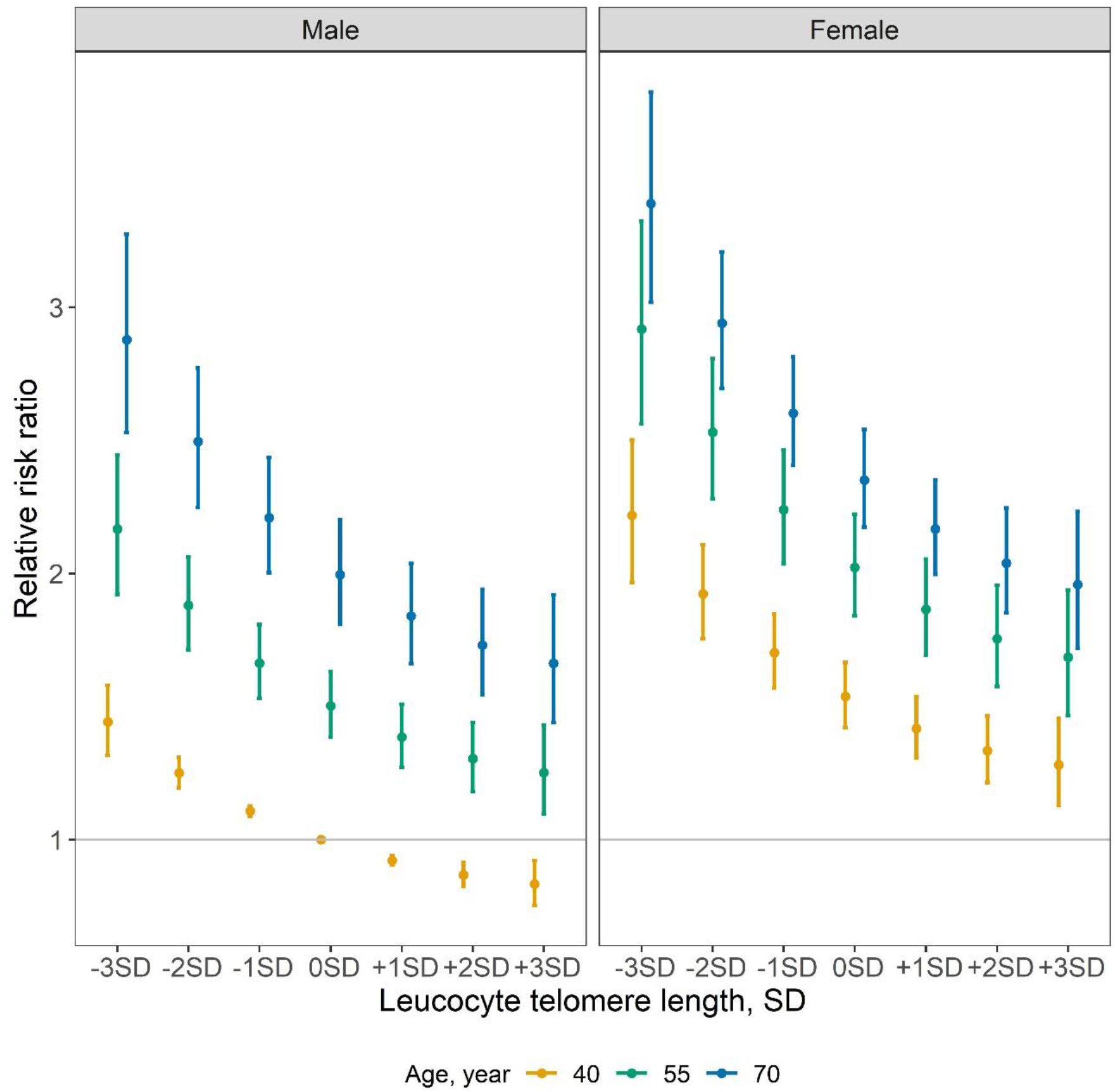
Relative risk ratios for the association of standardised leucocyte telomere length with frailty, by age and sex. **Legend:** Relative risk ratios derived from the estimates of age and leucocyte telomere length (LTL) for the frailty vs non–frailty model shown in Table 4, compared to a 40 year old male whilst holding all the rest of the covariates constant. The gradient in the association between age and LTL is shown, with a slightly sharper decline for participants with LTL <0SD compared to >0SD (p for LTL quadratic term=0.03) in both males and females. Compared to males, females have a higher risk of frailty in any given age or LTL group. Within sex, the age differences are more evident in males (p for interaction <0·0001).

**Table 4:**
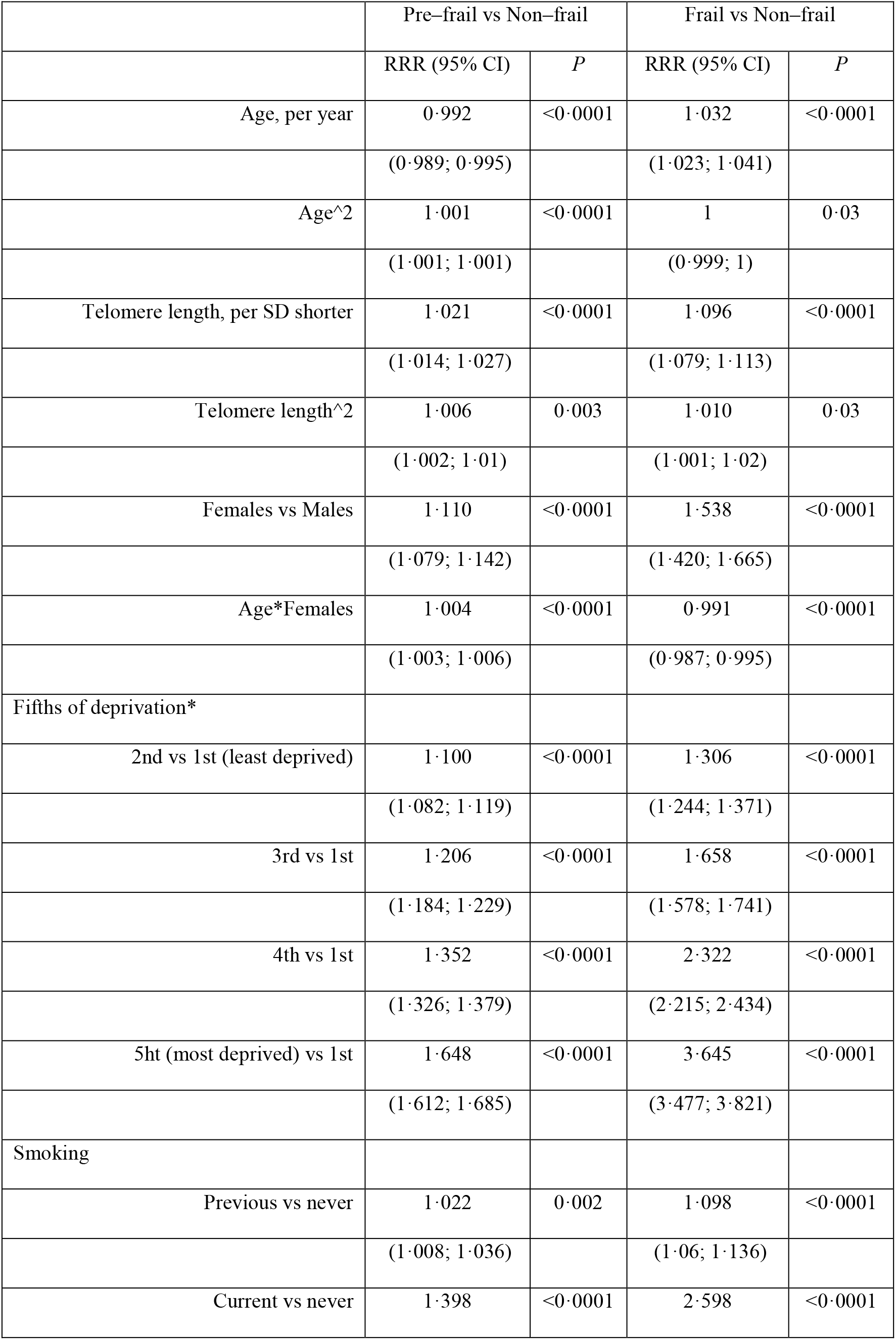
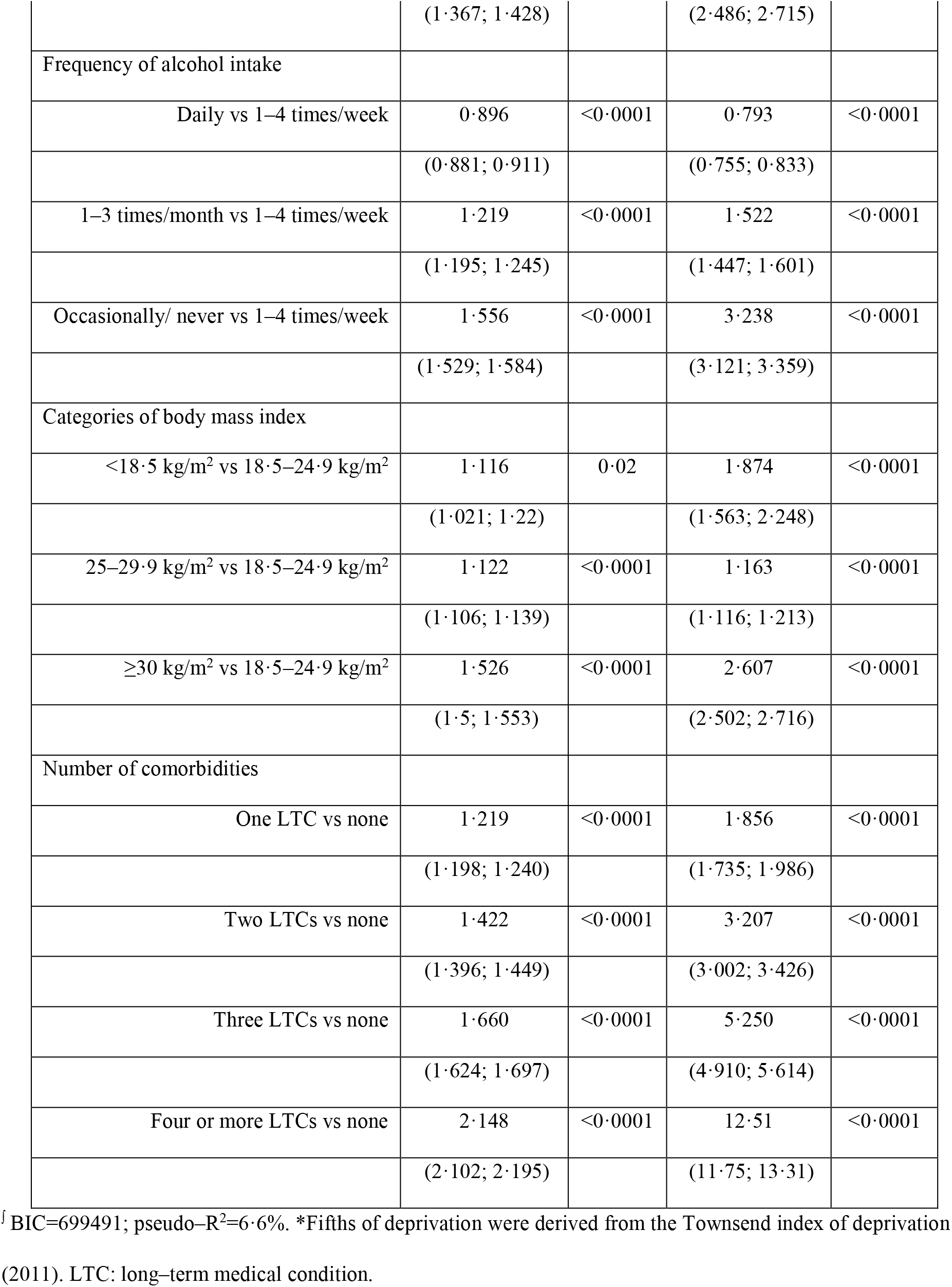
Adjusted relative risk ratios (RRR) and 95% confidence intervals (95% CI) from a multinomial logit model^∫^ on frailty.

|  | Pre-frail vs Non-frail |  | Frail vs Non-frail |  |
| --- | --- | --- | --- | --- |
|  | RRR (95% CI) | *P* | RRR (95% CI) | *P* |
| Age, per year | 0.992 | <0.0001 | 1.032 | <0.0001 |
|  | (0.989; 0.995) |  | (1.023; 1.041) |  |
| Age<sup>2</sup> | 1.001 | <0.0001 | 1 | 0.03 |
|  | (1.001; 1.001) |  | (0.999; 1) |  |
| Telomere length, per SD shorter | 1.021 | <0.0001 | 1.096 | <0.0001 |
|  | (1.014; 1.027) |  | (1.079; 1.113) |  |
| Telomere length<sup>2</sup> | 1.006 | 0.003 | 1.010 | 0.03 |
|  | (1.002; 1.01) |  | (1.001; 1.02) |  |
| Females vs Males | 1.110 | <0.0001 | 1.538 | <0.0001 |
|  | (1.079; 1.142) |  | (1.420; 1.665) |  |
| Age*Females | 1.004 | <0.0001 | 0.991 | <0.0001 |
|  | (1.003; 1.006) |  | (0.987; 0.995) |  |
| Fifths of deprivation* |  |  |  |  |
| 2nd vs 1st (least deprived) | 1.100 | <0.0001 | 1.306 | <0.0001 |
|  | (1.082; 1.119) |  | (1.244; 1.371) |  |
| 3rd vs 1st | 1.206 | <0.0001 | 1.658 | <0.0001 |
|  | (1.184; 1.229) |  | (1.578; 1.741) |  |
| 4th vs 1st | 1.352 | <0.0001 | 2.322 | <0.0001 |
|  | (1.326; 1.379) |  | (2.215; 2.434) |  |
| 5th (most deprived) vs 1st | 1.648 | <0.0001 | 3.645 | <0.0001 |
|  | (1.612; 1.685) |  | (3.477; 3.821) |  |
| Smoking |  |  |  |  |
| Previous vs never | 1.022 | 0.002 | 1.098 | <0.0001 |
|  | (1.008; 1.036) |  | (1.06; 1.136) |  |
| Current vs never | 1.398 | <0.0001 | 2.598 | <0.0001 |
|  | (1.367; 1.428) |  | (2.486; 2.715) |  |
| Frequency of alcohol intake |  |  |  |  |
| Daily vs 1–4 times/week | 0.896 | <0.0001 | 0.793 | <0.0001 |
|  | (0.881; 0.911) |  | (0.755; 0.833) |  |
| 1–3 times/month vs 1–4 times/week | 1.219 | <0.0001 | 1.522 | <0.0001 |
|  | (1.195; 1.245) |  | (1.447; 1.601) |  |
| Occasionally/ never vs 1–4 times/week | 1.556 | <0.0001 | 3.238 | <0.0001 |
|  | (1.529; 1.584) |  | (3.121; 3.359) |  |
| Categories of body mass index |  |  |  |  |
| <18.5 kg/m<sup>2</sup> vs 18.5–24.9 kg/m<sup>2</sup> | 1.116 | 0.02 | 1.874 | <0.0001 |
|  | (1.021; 1.22) |  | (1.563; 2.248) |  |
| 25–29.9 kg/m<sup>2</sup> vs 18.5–24.9 kg/m<sup>2</sup> | 1.122 | <0.0001 | 1.163 | <0.0001 |
|  | (1.106; 1.139) |  | (1.116; 1.213) |  |
| ≥30 kg/m<sup>2</sup> vs 18.5–24.9 kg/m<sup>2</sup> | 1.526 | <0.0001 | 2.607 | <0.0001 |
|  | (1.5; 1.553) |  | (2.502; 2.716) |  |
| Number of comorbidities |  |  |  |  |
| One LTC vs none | 1.219 | <0.0001 | 1.856 | <0.0001 |
|  | (1.198; 1.240) |  | (1.735; 1.986) |  |
| Two LTCs vs none | 1.422 | <0.0001 | 3.207 | <0.0001 |
|  | (1.396; 1.449) |  | (3.002; 3.426) |  |
| Three LTCs vs none | 1.660 | <0.0001 | 5.250 | <0.0001 |
|  | (1.624; 1.697) |  | (4.910; 5.614) |  |
| Four or more LTCs vs none | 2.148 | <0.0001 | 12.51 | <0.0001 |
|  | (2.102; 2.195) |  | (11.75; 13.31) |  |

<sup>f</sup> BIC=699491; pseudo-R<sup>2</sup>=6.6%. \*Fifths of deprivation were derived from the Townsend index of deprivation
(2011). LTC: long-term medical condition.

Correcting for regression dilution bias further strengthened the associations of LTL with both frailty conditions (3·1% (2·1%; 4·0%) and 14% (12%; 17%) higher, for pre–frailty and frailty respectively per one SD shorter LTL). Adjustment for the associations of LTL with 123 prevalent diseases reduced the associations of LTL with both pre–frailty and frailty but both remained significant (1·4% (0·8%; 2·1%) and 4·5% (2·9%; 6·1%) higher per one SD shorter LTL, p<0·0001 for both).

In the full model, sex and other established risk factors were substantially associated with both pre–frailty and frailty (**Table 4**). For example, compared with men, women had about 50% higher relative risk of being frail as opposed to being non–frail, while being in the highest fifth of social deprivation involved about 3·5–fold higher risk than being in the lowest fifth. Each additional LTC was associated with sharply higher risk of frailty. In particular, participants with at least four LTCs had about 12·5–fold higher risk of frailty compared with those with no LTC (**Table 4**). Overall, the variables we analysed explained approximately 6·6% of the variance in the distribution of the frailty phenotypes.

The predicted absolute frequency of frailty, derived from the full model, across the population distribution of age, stratified by LTL and accounting for other risk factors, is shown in **Figure 2**. In each age group, there was a gradient in increased frequency of frailty moving from longer to shorter LTL without any threshold effect. The strength of the association between LTL and frailty appeared similar in each of the different age categories. Thus, in each age group, the frequency of frailty was about 15% higher in participants with LTL one SD shorter vs one SD longer than the mean and about 33% higher between two SD either side of the mean (**Figure 2**).

**Figure 2:**
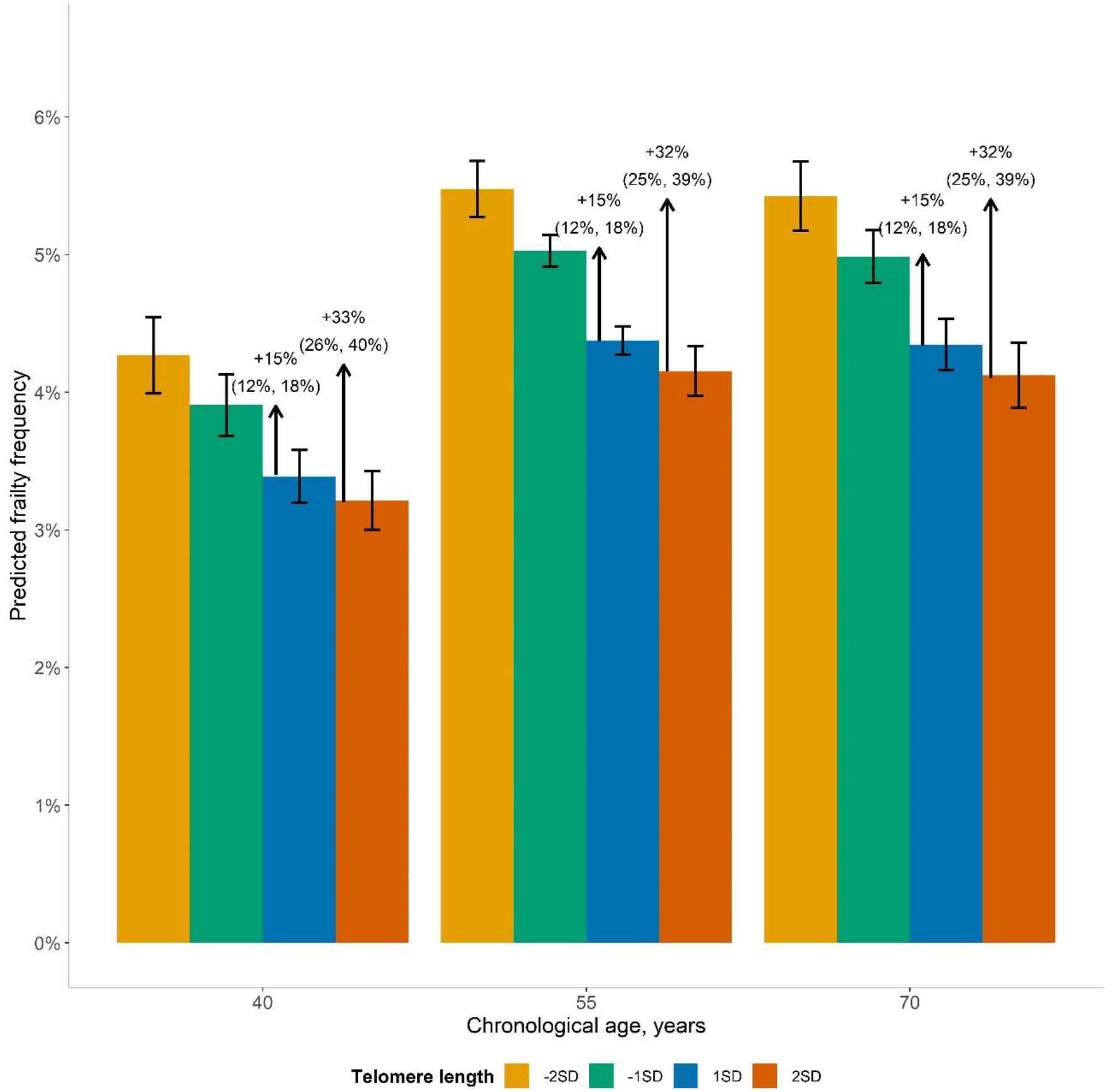
Predicted frequencies of frailty by chronological age, over four specific telomere lengths. **Legend:** Predicted frequency of frailty is derived from the estimates of age and leucocyte telomere length (LTL) for the frailty vs non–frailty model shown in Table 4, holding all the rest of the covariates at their observed values. Bars indicate the average predicted frequency of frailty, whilst error bars indicate the 95% confidence interval (CI). The ratio between two predicted frequencies (95% CI) is also given. There are approximately 2% (8,364) participants with LTL equal to -2SD or +2SD, and 9% (41,768) participants with LTL equal to -1SD or +1SD.

In the causal inference analysis there was a similar trend towards an association between shorter LTL and greater risk of frailty with a point estimate that overlapped with the observational association (**Supplementary Figure 4**). However, the 95% confidence intervals were wide and the association was not significant (OR 1·08 (0·98; 1·19) per one SD shorter genetically–determined LTL (p=0·13) from the MR–Median model). Different MR approaches (see **Supplementary Methods**) yielded similar results (**Supplementary Figure 4**); in particular, there was no evidence of substantial pleiotropy (MR Egger intercept’s p–value=0·60). Adjustment for additional covariates did not attenuate the trend (**Supplementary Figure 4**).

## Discussion

Utilising the powerful resource of the UK Biobank, in which we measured LTL in over 472,000 participants, we report a significant association between shorter LTL and greater risk of frailty in a contemporary population. We demonstrate that progressively shorter telomere lengths associate with non–frailty through pre–frailty to frailty across all age groups. The association between shorter LTL and greater risk of frailty remained significant after adjustment for other established determinants of frailty such as age, sex, alcohol intake, smoking, obesity, deprivation and multimorbidity. Furthermore, the associations of shorter LTL with pre–frailty and frailty were at least partly independent of the potential associations between variation in LTL and 123 prevalent diseases spanning multiple body systems.

A few studies have previously examined the relationship between frailty and telomere with variable findings. Two recent meta–analyses of these studies, including up to 3,268^14^ and 10,079^16^ individuals respectively, did not show a consistent association between shorter LTL and frailty indices. While several factors, including the age ranges and ethnicities studied, the method of telomere length measurement and definition of frailty, may contribute to the heterogeneous findings, the most likely reason is statistical power. Our study analysed over 40–fold more participants than the largest meta–analysis providing greater power to detect any association between variation in LTL and frailty as well as pre–frailty.

Fried’s frailty index^3^ integrates five different functional measures. We confirmed both an association of similar magnitudes of LTL with individual components of this index and also, importantly, that the association is stronger as the number of frailty indicators increase. These findings indicate that the observed association is not due to one of the component phenotypes.

At a tissue level, LTL is a determinant of replicative capacity and tissue repair,^10,13^ Thus, the association of shorter LTL with increased risk of frailty could be explained by earlier exhaustion of these functions across multiple body systems. However, a notable finding was that the *relative* increase in risk of frailty with shorter telomeres was similar in different age groups and did not increase with age. This suggests that LTL is not simply accelerating the effect of chronological age on risk of frailty as a biomarker of premature ageing. This is in accord with recent concepts around telomere dynamics and ageing–related diseases.^13^ However, it should be noted that the age range at recruitment of participants in UK was relatively narrow at between 40–69 years and we cannot exclude the possibility that the relative association of shorter LTL with risk of frailty might be stronger at older ages. A further concern about UKB is that, because recruitment was voluntary and required participants to travel to a recruitment centre, it may have recruited relatively “healthy” individuals particularly at older ages (60–69).^24^ This could impact on the prevalence of the frailty phenotype and hence the generalisability of the findings.^24^ However, we observed similar estimates for the age and sex–specific rates of frailty syndrome at the overlapping age range available (60–75 years) with two other studies available in the UK: those derived from the Hertfordshire Cohort Study^25^ and from the English Longitudinal Study of Ageing.^26^ Furthermore, previous analysis has shown that even if the prevalence of a disorder is different in UKB compared with a general population, it should not impact on its relative association with a risk factor.^27^

While the association of shorter LTL with risk of frailty was highly significant and there was approximately 33% higher risk of frailty in those with two SD shorter LTL compared with those with two SD longer LTL than average, the association needs to be viewed in context of other risk factors for frailty. As shown in Table 4, socio–demographic and lifestyle factors such as social deprivation, BMI, alcohol intake, smoking and presence of co–morbidity individually all had much more powerful associations than LTL. However, even taken in total the analysed factors only explained about 6·6% of the risk of frailty.

Although the richness of the information on participants in UKB allowed us to adjust for several relevant factors in assessing the association of LTL with risk of frailty, a cross–sectional analysis cannot infer causation. To investigate whether the association of LTL with risk if frailty was causal, we deployed Mendelian Randomisation using 131 genetic variants associated with LTL^12^ as instruments. Although this showed an association that was concordant with the observational finding both directionally and in terms of effect size, the 95% margins were wide, indicating limited power to confirm or exclude a genetic association. This probably reflects a combination of the low overall prevalence of frailty in the studied population and the relatively low strength of the genetic instruments, which explain less than 5% of the variation in LTL.^12^ Therefore, we cannot exclude the possibility that the observed association between shorter LTL and increased risk of frailty is due to residual confounding which we have not accounted for.

More broadly, our analysis and findings have relevance to definitional approaches, which have been developed for the characterisation of frailty.^2,3^ The concept of frailty attempts to explain the heterogeneity in health and functional status, as individuals get older, which is thought to arise from a reduction in reserve capacity in various physiologic systems. In the present analysis, LTL–frailty associations were at least partly independent of the presence of long–term comorbidities, included either as number of conditions or via the residuals derived from LTL regressed on the wider set of 123 morbidities. This suggests that there is, in addition to “accumulated morbidity”, an element of the frailty syndrome that is independent of comorbidities, as proposed by Fried et al.^3^ thereby supporting the notion that the syndromic approach may identify elements of vulnerability and resilience that distinguish frailty from disability or disease that accumulate over time.^1,28^

Despite the scale of our study and the uniform and detailed phenotypic characterisation in UKB, some limitations, in addition to those discussed earlier, should be considered in the interpretation of our findings. First, as information to derive the frailty phenotype was only collected at baseline, we are unable to investigate any relationship of inter–individual variation in LTL to future development of frailty. Similarly, our single point estimate of LTL precludes analysis of the association of any changes in LTL with age and development of frailty. Finally, UKB predominantly comprises individuals of white ethnicity. There are differences in average LTL (adjusting for age and gender) in participants from different ethnicities^17^ and whether the association of LTL with risk of frailty differs in participants from different backgrounds remains to be investigated.

In summary, we show that shorter LTL is associated with greater risk of syndromic frailty and that this association is independent of other risk factors but partly explained by the causal association of LTL with diseases across multiple body systems. Our findings provide evidence for an additional biological factor associated with the risk of frailty.

## Data Availability

The MTA with UK Biobank precludes direct data sharing. All derived data are deposited back to UK Biobank and are available upon request to UK Biobank. Dissemination of the findings to study participants will be through the UK Biobank website.

## Contributions

VB, CPN, VC and NJS conceived the project and analysis plan. VB, QW and CM assembled the data. VB, CPN and VC have verified the underlying data. VB undertook the majority of the analysis with suggestions and input from all the co–authors. VB undertook the literature review. VB, CPN, VC, EC, NH, CC and NJS drafted the manuscript and all authors revised it. VC, CPN, JRT, JND and NJS (Principal investigator) secured funding for the telomere length measurements. All authors approved the manuscript and the decision to submit the paper for publication.

## Acknowledgments

University of Leicester investigators are supported by the National Institute for Health Research (NIHR) Leicester Cardiovascular Biomedical Research Centre (BRC–1215–20010). CPN is funded by the BHF (SP/16/4/32697). Cambridge University investigators are supported by the BHF (RG/13/13/30194; RG/18/13/33946), Health Data Research UK, NIHR Cambridge Biomedical Research Centre (BRC–1215–20014), NIHR Blood and Transplant Research Unit in Donor Health and Genomics (NIHR BTRU–2014–10024) and MRC (MR/L003120/1). JD holds a BHF Personal Professorship and NIHR Senior Investigator Award. AMW and EA received support from the EU/EFPIA Innovative Medicines Initiative Joint Undertaking BigData@Heart (11607). University of Southampton investigators are supported by Medical Research Council (MRC) [4050502589 (MRC LEU)], National Institute for Health Research (NIHR) Southampton Biomedical Research Centre, University of Southampton and University Hospital Southampton NHS Foundation Trust, and NIHR Oxford Biomedical Research Centre, University of Oxford.

