## Supplementary material for "Association of leucocyte telomere length with frailty: a large–scale cross–sectional analysis in UK Biobank"

### Online supplementary material

#### Methods

*Statistical analysis*

Seven sequential multinomial logistic regression models were initially tested: i) a univariable model including only age (M1a), ii) a univariable model including only leucocyte telomere length (LTL; M1b), iii) a model including age, LTL and their interaction (M2), iv) M2 with an additional quadratic term for age (M3a), v) M2 with an additional quadratic term for LTL (M3b), vi) a model with age, LTL, their interaction and their quadratic terms (M4), and vii) a model with age, LTL and their quadratic terms (M5). Models were compared using the Bayesian information criterion (BIC), and the model with the lowest BIC was selected.

Following the best model selection for age and LTL (M5), a model including sex (females/ males) as an additional predictor and further models with interaction terms of sex with age and LTL were also evaluated. Finally, the best selected model from this step (M5+sex+age*sex) was additionally adjusted for other well–known determinants of frailty (see main paper).

In a sensitivity analysis we also performed a generalised ordinal model,[1] loosening the assumption of proportionality, with the frailty phenotype (none/ pre–frail/ frail) as an outcome. In addition, we performed binary logistic regression models to investigate the association between LTL and the individual frailty indicators, as well a multinomial logistic regression model for the association between LTL and the total number of frailty indicators.

To explore whether LTL is causally associated with the phenotype of frailty we performed a genetic analysis using131 genetic variants associated with LTL[2] as instrumental variables. To estimate the genetic associations of the variants with z–standardised LTL (beta1) we conducted linear regression analysis between the z–standardised LTL and each genetic variant while adjusting for age (centred at the age of 40), sex, array, and the first 10 principal components as described previously.[2] To estimate the associations of genetic variants with frailty (beta2) we performed two different logistic regression analyses between frailty and each genetic variant in turn. In the first analysis we adjusted the model only for the same covariates listed above, whilst in the second analysis we additionally adjusted the model deprivation, alcohol intake, smoking, BMI and LCTs, as these were significantly associated with the prevalence of frailty using the observational data. The causal association between LTL and the phenotype of frailty was assessed through one–sample Mendelian Randomization (MR),[3] utilising different methods to test for robustness of findings. In particular, to estimate the odds ratio for frailty per one SD shorter LTL we used four different MR functions: i) the inverse variance weighted MR (IVW), ii) the median based estimator (Median) and iii) the maximum likelihood (MaxLik) of the "MendelianRandomisation" package,[4] and iv) the Robust Adjusted Profile (RAPS) of the "mr.raps" package.[5] The selected methods complement each other in the assumptions that should be satisfied in order to produce valid results and were used in parallel. Particularly, the IVW is the most efficient and robust method in the absence of pleiotropy, the Median is additionally robust in the presence of outliers, the MaxLik allows a) for uncertainty in the genetic associations with the exposure and b) for genetic associations with the exposure and with the outcome for each variant to be correlated and the RAPS method overcomes challenges related to measurement error, weak or invalid (due to pleiotropy) measurements and selection bias (due to weak instrument). Therefore a combination of these methods provides the best evidence for the presence of a causal association.[6,7] In addition, the presence of pleiotropy was assessed through the MR–Egger analysis.[8] MR analyses were performed in R version 3·1·6,[12] whilst the regression models were performed in Stata v16·0.[13] The significance level for all associations was set at 0·05.

**Supplementary Table 1: Adjusted relative risk ratios (RRR) and 95% confidence intervals (95% CI) from a multinomial regression model[ʃ] of age, sex and leucocyte telomere length on number of frailty indicators.**

| | Number of frailty indicators | | | | | | | | | |
|---|---|---|---|---|---|---|---|---|---|---|
| | 1 vs 0 | | 2 vs 0 | | 3 vs 0 | | 4 vs 0 | | 5 vs 0 | |
| | RRR (95% CI) | *P* | RRR (95% CI) | *P* | RRR (95% CI) | *P* | RRR (95% CI) | *P* | RRR (95% CI) | *P* |
| Age, per year increase | 0·99 | <0·0001 | 1·004 | 0·17 | 1·03 | <0·0001 | 1·04 | <0·0001 | 1·11 | <0·0001 |
| | (0·985; 0·992) | | (0·998; 1·009) | | (1·02; 1·04) | | (1·02; 1·06) | | (1·06; 1·17) | |
| Age^2 | 1·001 | <0·0001 | 1 | <0·0001 | 1 | 0·34 | 0·999 | 0·03 | 0·998 | 0·002 |
| | (1·001; 1·001) | | (1; 1·001) | | (1; 1) | | (0·999; 1) | | (0·996; 0·999) | |
| Telomere length, per SD shorter | 1·01 | <0·0001 | 1·04 | <0·0001 | 1·09 | <0·0001 | 1·12 | <0·0001 | 1·21 | <0·0001 |
| | (1·007; 1·02) | | (1·03; 1·05) | | (1·07; 1·11) | | (1·09; 1·16) | | (1·11; 1·32) | |
| Telomere length^2 | 1·002 | 0·29 | 1·02 | <0·0001 | 1·02 | 0·003 | 1 | 0·98 | 0·97 | 0·28 |
| | (0·998; 1·01) | | (1·011; 1·02) | | (1·005; 1·03) | | (0·981; 1·02) | | (0·92; 1·02) | |
| Females vs Males | 1·07 | <0·0001 | 1·27 | <0·0001 | 1·54 | <0·0001 | 1·61 | <0·0001 | 2·03 | 0·003 |
| | (1·04; 1·1) | | (1·21; 1·34) | | (1·4; 1·68) | | (1·36; 1·9) | | (1·28; 3·21) | |
| Age*Females | 1·005 | <0·0001 | 1 | 0·86 | 0·993 | 0·001 | 0·986 | 0·001 | 0·972 | 0·02 |
| | (1·003; 1·007) | | (0·998; 1·003) | | (0·988; 0·997) | | (0·978; 0·995) | | (0·95; 0·995) | |

[ʃ]Model additionally adjusted for fifths of Townsend index of deprivation (2011), smoking, alcohol intake, body mass index and number of long–term medical conditions.

**Supplementary Table 2: Relative risk ratios (RRR) and 95% confidence intervals (95% CI) from multinomial logit models of age and leucocyte telomere length on frailty.**

| | BIC; | Pre–frail *vs.* Non–frail | | Frail *vs.* Non–frail | |
|---|---|---|---|---|---|
| | pseudo–$R^2$ | RRR (95% CI) | *P* | RRR (95% CI) | *P* |
| *M1a* | 745093; | | | | |
| | 0·45% | | | | |
| Age, per year | | 1·019 | <0·0001 | 1·037 | <0·0001 |
| | | (1·018; 1·02) | | (1·035; 1·039) | |
| *M1b* | 747826; | | | | |
| | 0·09% | | | | |
| Telomere length, per SD shorter | | 1·051 | <0·0001 | 1·178 | <0·0001 |
| | | (1·045; 1·058) | | (1·161; 1·195) | |
| *M2* | 744886; | | | | |
| | 0·49% | | | | |
| Age, per year | | 1·019 | <0·0001 | 1·034 | <0·0001 |
| | | (1·018; 1·02) | | (1·032; 1·036) | |
| Telomere length, per SD shorter | | 0·988 | 0·09 | 1·083 | <0·0001 |
| | | (0·974; 1·002) | | (1·044; 1·124) | |
| Age*Telomere length | | 1·002 | <0·0001 | 1·002 | 0·05 |
| | | (1·001; 1·003) | | (1; 1·004) | |
| *M3a* | 744663; | | | | |
| | 0·52% | | | | |
| Age, per year | | 0·994 | 0·001 | 1·035 | <0·0001 |
| | | (0·991; 0·998) | | (1·027; 1·044) | |
| Age^2 | | 1·001 | <0·0001 | 1 | 0·98 |
| | | (1·001; 1·001) | | (1; 1) | |
| Telomere length, per SD shorter | | 1·007 | 0·36 | 1·081 | <0·0001 |
| | | (0·992; 1·021) | | (1·041; 1·122) | |
| Age*Telomere length | | 1·001 | 0·04 | 1·002 | 0·05 |
| | | (1; 1·002) | | (1; 1·004) | |
| *M3b* | 744894; | | | | |
| | 0·49% | | | | |
| Age, per year | | 1·019 | <0·0001 | 1·034 | <0·0001 |
| | | (1·018; 1·020) | | (1·032; 1·036) | |
| Telomere length, per SD shorter | | 0·993 | 0·37 | 1·094 | <0·0001 |
| | | (0·979; 1·008) | | (1·054; 1·134) | |
| Telomere length^2 | | 1·002 | 0·001 | 1·001 | 0·002 |
| | | (1·001; 1·002) | | (0·999; 1·003) | |
| Age*Telomere length | | 1·007 | <0·0001 | 1·015 | 0·23 |
| | | (1·003; 1·011) | | (1·005; 1·024) | |
| *M4* | 744669; | | | | |
| | 0·52% | | | | |
| Age, per year | | 0·994 | <0·0001 | 1·035 | <0·0001 |
| | | (0·991; 0·997) | | (1·027; 1·043) | |
| Age^2 | | 1·001 | <0·0001 | 1 | 0·97 |
| | | (1·001; 1·001) | | (1; 1) | |
| Telomere length, per SD shorter | | 1·013 | 0·08 | 1·092 | <0·0001 |

| | | | | | |
|---|---|---|---|---|---|
| | | (0·998; 1·028) | | (1·051; 1·134) | |
| Telomere length^2 | | 1·008 | <0·0001 | 1·015 | 0·002 |
| | | (1·004; 1·012) | | (1·005; 1·024) | |
| Age*Telomere length | | 1 | 0·27 | 1·001 | 0·23 |
| | | (1; 1·001) | | (0·999; 1·003) | |
| ***M5*** | 744645; | | | | |
| | 0·52% | | | | |
| Age, per year | | 0·994 | <0·0001 | 1·034 | <0·0001 |
| | | (0·991; 0·997) | | (1·026; 1·042) | |
| Age^2 | | 1·001 | <0·0001 | 1 | 0·78 |
| | | (1·001; 1·001) | | (1; 1) | |
| Telomere length, per SD shorter | | 1·02 | <0·0001 | 1·115 | <0·0001 |
| | | (1·014; 1·027) | | (1·098; 1·131) | |
| Telomere length^2 | | 1·008 | <0·0001 | 1·016 | <0·0001 |
| | | (1·004; 1·012) | | (1·007; 1·025) | |

The model in **bold** indicates the best model selected on the basis of minimizing the BIC.

**Supplementary Table 3: Adjusted odds ratios (OR) and 95% confidence intervals (95% CI) from a generalised ordinal model of age and leucocyte telomere length on frailty.**

| | Pre–/ Frail *vs.* Non–frail | | Frail *vs.* Pre– and Non–frail | |
|---|---|---|---|---|
| | OR (95% CI) | *P* | OR (95% CI) | *P* |
| *M5* | | | | |
| Age, per year | 0·997 | 0·05 | 1·039 | <0·0001 |
| | (0·994; 1·000) | | (1·031; 1·047) | |
| Age^2 | 1·001 | <0·0001 | 1 | 0·001 |
| | (1·001; 1·001) | | (0·999; 1) | |
| Telomere length, per SD shorter | 1·029 | <0·0001 | 1·104 | <0·0001 |
| | (1·023; 1·035) | | (1·089; 1·12) | |
| Telomere length^2 | 1·010 | <0·0001 | 1·010 | <0·0001 |
| | (1·006; 1·013) | | (1·006; 1·013) | |

BIC=744629; pseudo–$R^2$=0·52%.

**Supplementary Table 4: Adjusted relative risk ratios (RRR) and 95% confidence intervals (95% CI) from multinomial logit models of age, leucocyte telomere length (LTL) and sex on frailty.**

| | BIC; | Pre–frail vs Non–frail | | Frail vs Non–frail | |
|---|---|---|---|---|---|
| | pseudo–$R^2$ | RRR (95% CI) | *P* | RRR (95% CI) | *P* |
| *M5+sex* | 743217; | | | | |
| | 0·72% | | | | |
| Age, per year | | 0·993 | <0·0001 | 1·031 | <0·0001 |
| | | (0·989; 0·996) | | (1·023; 1·039) | |
| Age^2 | | 1·001 | <0·0001 | 1 | 0·26 |
| | | (1·001; 1·001) | | (1; 1) | |
| Telomere length, per SD shorter | | 1·030 | <0·0001 | 1·136 | <0·0001 |
| | | (1·023; 1·036) | | (1·119; 1·153) | |
| Telomere length^2 | | 1·008 | <0·0001 | 1·016 | 0·001 |
| | | (1·004; 1·012) | | (1·007; 1·025) | |
| Female vs male | | 1·219 | <0·0001 | 1·499 | <0·0001 |
| | | (1·204; 1·234) | | (1·455; 1·543) | |
| ***M5+sex+age*sex*** | 743168; | | | | |
| | 0·73% | | | | |
| Age, per year | | 0·989 | <0·0001 | 1·034 | <0·0001 |
| | | (0·986; 0·992) | | (1·026; 1·043) | |
| Age^2 | | 1·001 | <0·0001 | 1 | 0·29 |
| | | (1·001; 1·001) | | (1; 1) | |
| Telomere length, per SD shorter | | 1·030 | <0·0001 | 1·136 | <0·0001 |
| | | (1·024; 1·037) | | (1·119; 1·153) | |
| Telomere length^2 | | 1·009 | <0·0001 | 1·016 | 0·001 |
| | | (1·005; 1·012) | | (1·007; 1·024) | |
| Females vs Males | | 1·103 | <0·0001 | 1·629 | <0·0001 |
| | | (1·072; 1·134) | | (1·511; 1·757) | |
| Age*Females | | 1·006 | <0·0001 | 0·996 | 0·03 |
| | | (1·005; 1·008) | | (0·992; 1·000) | |
| *M5+sex+age*sex+LTL*sex* | 743188; | | | | |
| | 0·73% | | | | |
| Age, per year | | 0·989 | <0·0001 | 1·034 | <0·0001 |
| | | (0·986; 0·992) | | (1·025; 1·042) | |
| Age^2 | | 1·001 | <0·0001 | 1 | 0·28 |
| | | (1·001; 1·001) | | (1; 1) | |
| Telomere length, per SD shorter | | 1·027 | <0·0001 | 1·158 | <0·0001 |
| | | (1·018; 1·037) | | (1·131; 1·186) | |
| Telomere length^2 | | 1·009 | <0·0001 | 1·014 | 0·002 |
| | | (1·005; 1·013) | | (1·005; 1·023) | |
| Females vs Males | | 1·105 | <0·0001 | 1·615 | <0·0001 |
| | | (1·074; 1·136) | | (1·497; 1·743) | |
| Age*Females | | 1·006 | <0·0001 | 0·997 | 0·08 |
| | | (1·004; 1·007) | | (0·993; 1) | |
| Telomere length*Females | | 1·006 | 0·38 | 0·969 | 0·04 |
| | | (0·993; 1·019) | | (0·94; 0·999) | |

The model in **bold** indicates the best model selected on the basis of minimizing the BIC.

**Supplementary Figure 1: Flow of participants in the study.**

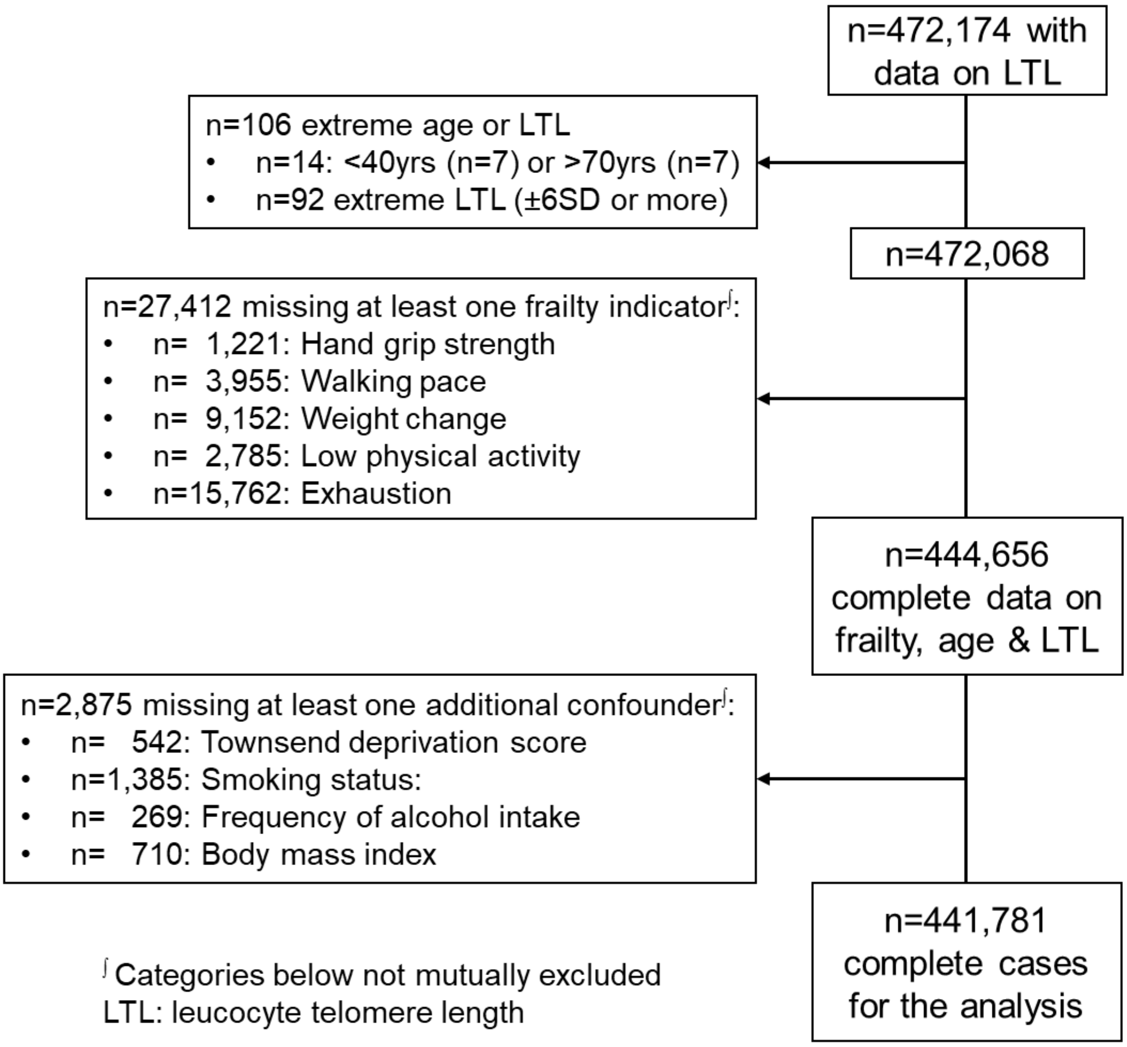

**Supplementary Figure 2: Participants' distribution by frailty status over age, as occurred from the observed data and the fitted values from model M5 (~age+age$^2$+LTL+LTL$^2$), for average LTL (0SD).**

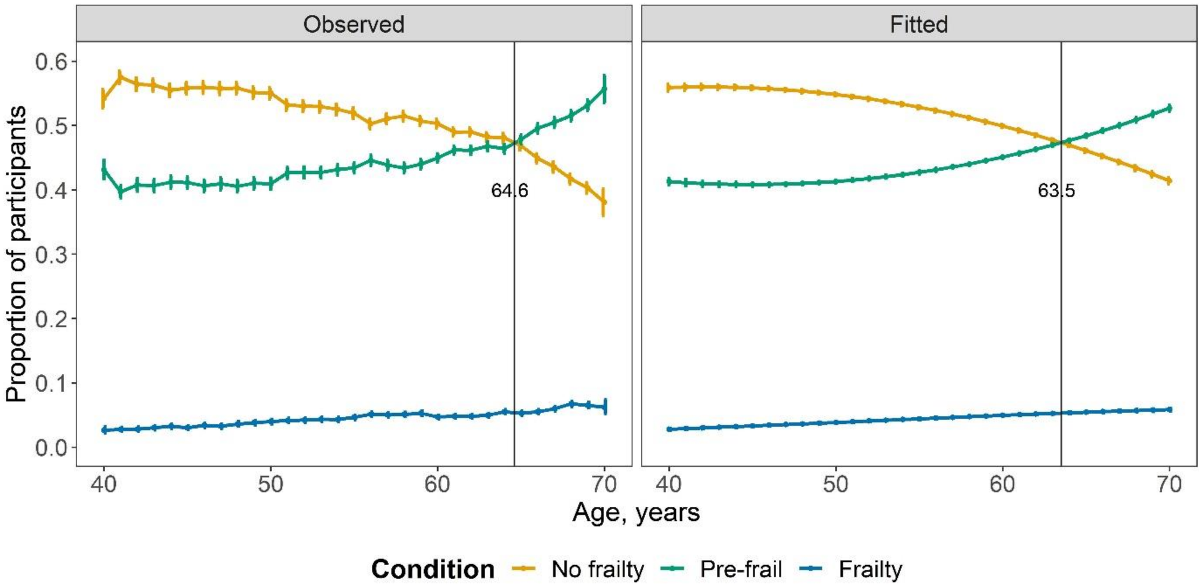

**Supplementary Figure 3: Results (odds ratios (95% confidence intervals)) from mendelian randomization (MR) and multinomial regression to assess respectively the causal (*MR*) and the observational (*Data*) association between telomere length and frailty.**

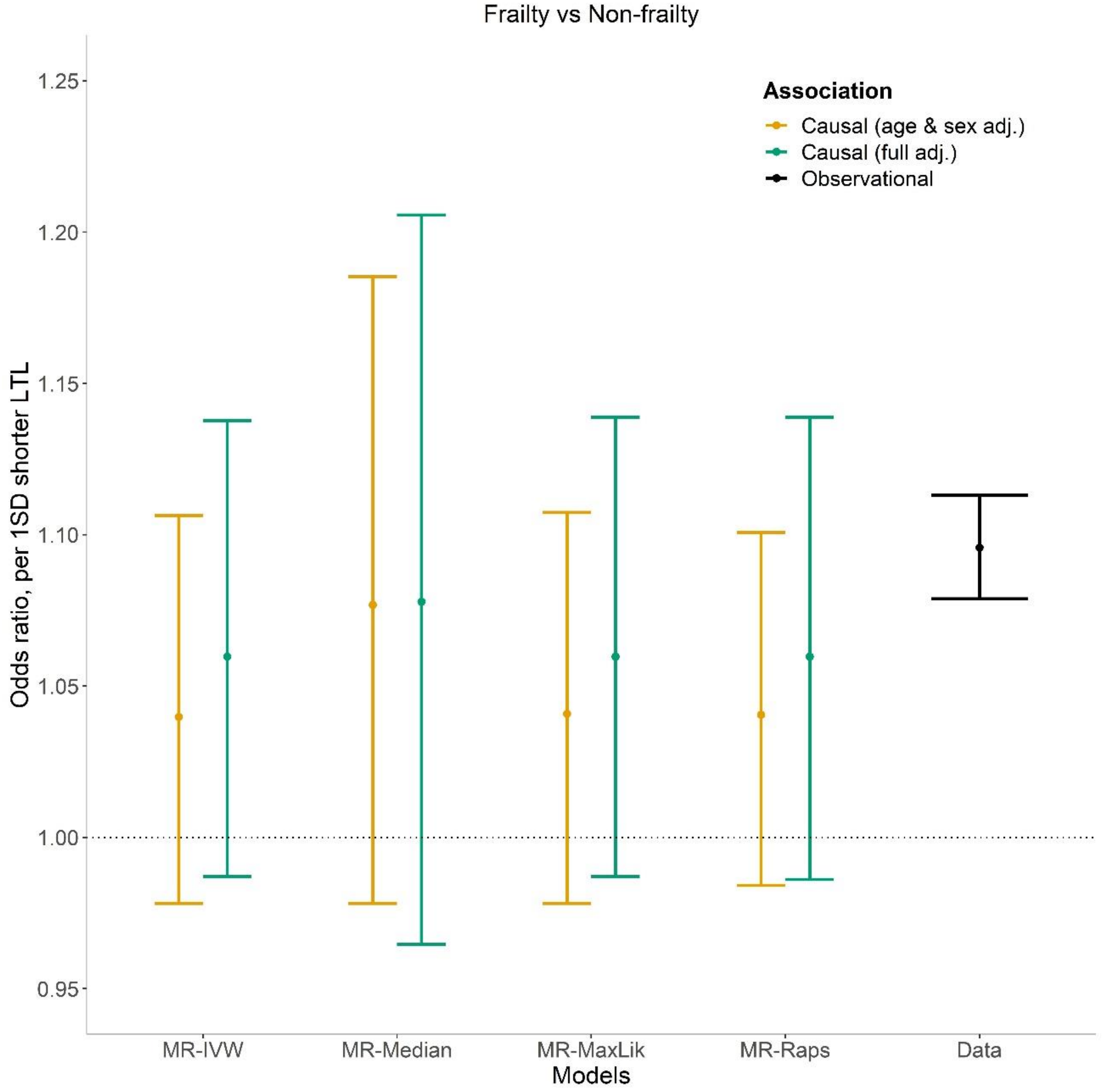


**Legend:** In addition to array and first 10 principal components of the age & sex adjusted causal model, covariates included in the fully adjusted causal model were deprivation, alcohol intake, smoking, body mass index, and number of long–term medical conditions.
